# The Cognitive-Functional Composite is sensitive to clinical progression in early dementia: longitudinal findings from the Catch-Cog study cohort

**DOI:** 10.1101/2020.02.06.20020859

**Authors:** Roos J. Jutten, John E. Harrison, A. Brunner, R. Vreeswijk, R.A.J. van Deelen, Frank Jan de Jong, Esther M. Opmeer, Craig W. Ritchie, André Aleman, Philip Scheltens, Sietske A.M. Sikkes

## Abstract

**Introduction:** In an attempt to capture clinically meaningful cognitive decline in early dementia, we developed the Cognitive-Functional Composite (CFC). We investigated the CFC’s sensitivity to decline in comparison to traditional clinical endpoints.

**Methods:** This longitudinal construct validation study included 148 participants with subjective cognitive decline, mild cognitive impairment or mild dementia. The CFC and traditional tests were administered at baseline, 3, 6 and 12 months. Sensitivity to change was investigated using linear mixed models and *r*^2^ effect-sizes.

**Results:** CFC scores declined over time (*β*=−.16,*p<*.*001*), with steepest decline observed in mild Alzheimer’s dementia (*β*=−.25,*p<*.*001*). The CFC showed medium-to-large effect-sizes at succeeding follow-up points (*r*^2^=.08–.42), exhibiting greater change than the Clinical Dementia Rating scale (*r*^2^=.02–.12). Moreover, change on the CFC was significantly associated with informant reports of cognitive decline (*β*=.38,*p*<.001).

**Discussion:** By showing sensitivity to decline, the CFC could enhance the monitoring of disease progression in dementia research and clinical practice.

## 1. Introduction

As Alzheimer’s disease (AD) clinical trials are increasingly targeting earlier disease stages [1], it is of crucial importance that outcome measures of efficacy are adapted to these novel target populations [2]. Common guidance for the selection of acceptable endpoints for use in clinical trials, as well as observational studies in general, holds that selected measures must exhibit acceptable levels of reliability, validity and sensitivity to change in the target population [3]. Furthermore, these tests must be free of range restrictions, and ideally be brief so as to avoid fatigue or ennui effects, as well as appropriate for cross-cultural use [4].

Whilst commonly employed cognitive and functional measures selected for AD clinical trials have tended to demonstrate acceptable levels of reliability, they have fared less well with regard to their validity and sensitivity to change over time. For example, floor- and ceiling effects in scoring have been observed on parts of the Alzheimer’s disease assessment scale – Cognitive subscale (ADAS-Cog [5]) when administered in mild cognitive impairment (MCI) and mild dementia, limiting its sensitivity to change over time [6-8]. Moreover, it has been questioned to what extent change on the ADAS-Cog, as well as other cognitive tests, reflect clinically meaningful changes [9, 10]. Challenges regarding validity have also been encountered when measuring functional skills in individuals with MCI or mild dementia, as most existing measures do not feature complex instrument activities of daily living (IADL), which are most prone to early cognitive decline, or items indexing contemporary everyday activities such as electronic banking and self-organized travel arrangements [11-13].

In the past few years the AD research community has sought to identify novel methods that yield a single, unitary and valid measure of efficacy, including elements of both cognitive and functional performance [14]. This has also been encouraged by the Food and Drug Administration (FDA) and European Medicine Agency (EMA) in their guidelines for AD drug trials [15, 16]. A measure that has found favor as a combined measure is the Clinical Dementia Rating (CDR) scale [17], particular its sum of boxes (CDR-SB) scoring [18, 19]. However, a challenge when employing the CDR has been the modest rate of change over time when the scale is employed in those living with the very earliest manifestations of the disease [20]. Further challenges have been that the CDR-SB scoring requires extensive training, is subject to variability among ethnicity and languages and showed only modest inter-rater reliability [21].

To fulfil the need for a reliable, valid, sensitive and clinically meaningful measure of cognitive decline in early clinical stages of AD, the Cognitive-Functional Composite (CFC) has been designed [22]. The CFC yields a brief measure (20-25 minutes) of both cognition and function, comprising seven existing cognitive tests focusing on memory and executive functioning [23] and the Amsterdam IADL Questionnaire (A-IADL-Q) [24-26]. We have previously demonstrated good psychometric qualities of the CFC, such as good test-retest reliability, feasibility of use, validity and quality for the target population [27, 28]. The current study aimed to investigate the sensitivity to change of the CFC in individuals with MCI and mild AD dementia over a period of one year, as well as performance of the same individuals over the same period on the CDR-SB [18], ADAS-Cog [5], and Alzheimer’s Disease Cooperative Study – Activities of Daily Living (ADCS-ADL) scale [29]. Second, we explored whether the CFC could be of use to capture change in individuals with subjective cognitive impairment (SCD), and Dementia with Lewy Bodies (DLB) which is the second common cause of dementia. We also sought to determine the clinical meaningfulness of decline detected by the CFC, by associating change on the CFC with informant-reports of decline in everyday abilities.

## 2. Methods

### 2.1. Study design and participants

In this longitudinal analysis, we employed data from the Capturing Change in Cognition (Catch-Cog) study: an international, observational cohort-study with baseline, 3, 6, and 12 month assessments [22]. Participants (N=173) and their study partners were included at the 1) Alzheimer Center Amsterdam, Amsterdam UMC, the Netherlands (AC, n=102); 2) Alzheimer Center of the Erasmus Medical Center, Rotterdam, the Netherlands (EMC, n=14); 3) University Medical Center Groningen, the Netherlands (UMCG, n=39); or 4) the Centre for Dementia Prevention, Edinburgh, Scotland (EDI, n=18). Before inclusion, participants had undergone a standard diagnostic work-up in their memory clinic, including at least medical history, neurological and neuropsychological examination. A subset of participants included in the AC had AD biomarkers available as measured by a cerebrospinal fluid lumbar puncture. Amyloid positivity was based on amyloid-*β* 1-42 values (cut-off ≤ 813 pg./ml [30]). In all centers, clinical diagnoses were made in a multidisciplinary consensus meeting including at least a neurologist, psychiatrist and neuropsychologist. Additionally in the UMCG, participants were recruited via advertisements in local newspapers. Individuals willing to participate were screened by a neuropsychologist and neurologist to investigate whether they were eligible for the current study.

Participants were included in the Catch-Cog study when they met the research criteria for subjective cognitive decline (SCD) [31], or the clinical criteria for MCI [32], possible or probable AD dementia [33], or possible or probable DLB dementia [22]. Other inclusion criteria were: 1) Mini-Mental State Examination (MMSE) score ≥ 18 [34]; 2) age ≥ 50; and 3) availability of a study partner who was able to understand the study information and willing to participate. Exclusion criteria were 1) presence of another neurological disorder than AD or DLB; 2) presence of a major psychiatric disorder such as severe personality disorder or depression (Geriatric Depression Scale score ≥ 6 [35]); 3) current abuse of alcohol or drugs; 4) simultaneously participating in a clinical trial.

Data were collected between October 2016 and December 2018. The Medical-Ethical Committee of the VU University Medical Center approved the study for all Dutch centers. The South East Scotland Research Ethic Committee approved the study for the Scottish site. All participants and their study partners provided written and oral informed consent.

### 2.2. The Cognitive-Functional Composite

Full details on the selected CFC measures have been reported elsewhere [22, 28]. Briefly, the cognitive test battery of the CFC includes the three ADAS-cog memory subscales Word Recognition (score range 0-12), Word Recall (score range 0-10) and Orientation (score range 0-8) [5]; the Controlled Oral Word Association Test (COWAT; letters D-A-T in Dutch and F-A-S in English, 60 seconds) [36]; Category Fluency Test (CFT; animals, 60 seconds) [36]; Digit Span Backward (DSB, score range 0-14) [37] [1] and Digit Symbol Substitution Test (DSST, 90 seconds) [38]. The functional component comprises the short version of the A-IADL-Q, a computerized, informant-based questionnaire consisting of 30 items covering a broad range of complex IADL [26]. Example items include cooking, managing finances and modern activities such as applying everyday technology [24, 25]. For each item, difficulty in performance is rated on a 5-point Likert scale (ranging from ‘no difficulty in performing this task’ to ‘no longer able to perform this task’). Scoring is performed using item response theory (IRT), resulting in a latent trait score (z-score) reflecting one’s IADL functioning [25].

To create CFC scores, the directionality of the three ADAS-Cog subtest scores is reversed so that higher scores reflected better performance. Subsequently, all cognitive subtest scores are z-transformed using baseline total group means and standard deviations (SD). The cognitive composite is computed as a weighted z-score of all seven cognitive subtests if at least five tests are available, with all available tests being equally weighted. This approach was chosen as it was previously shown to provide a reliable scoring method for this cognitive composite [23]. The functional component score is the A-IADL-Q latent trait z-score [25]. The overall CFC score is computed as an equally weighted z-score of the cognitive composite and A-IADL-Q scores, with higher scores indicating better performance. We previously showed that this scoring method results in a valid CFC score that is in line with clinical manifestations of different diagnostic groups and not affected by range restrictions in scoring [28].

### 2.3. Reference measures

As previously reported [22], commonly employed AD clinical trial measures were administered, including the ADAS-Cog-13 (total score range 0-85) [5], ADCS-ADL (total score range 0-78) [29], and the CDR-SB (total score range 0-18) [18]. The study partner version of the Cognitive Function Instrument (CFI) was administered as anchor measure of clinical decline. The CFI includes 14 items that enquire about decline in day-to-day cognitive and functional abilities, compared with one year ago (score range 0-14, higher scores reflecting more decline) [39]. It was originally developed to track decline in preclinical stages of AD, but the study partner version was also found to be useful to assess decline individuals with subtle cognitive impairment [40].

### 2.4. Procedures

Study visits took place at the hospital or the participant’s home, depending on the participant’s preference. At baseline, a number of 74 participants (43%) chose testing at home, and we aimed to keep assessment location constant within participants over time. CFC and traditional tests were administered at each follow-up time-point, so that head-to-head comparisons could be made between all measures. A trained rater administered the cognitive tests according to standardized instructions, starting with the MMSE and followed by the cognitive part of the CFC and the remaining ADAS-Cog-13 tests. In the meantime, the study partner completed the A-IADL-Q and CFI independently on an iPad. Finally, the rater completed the ADCS-ADL and CDR interview with the study partner. The content of each study visit was similar, except that existing, validated parallel versions were used for Word Recognition and Word Recall, specifically List 1, 2, 4 and 5 [5].

A shortened protocol was used in the SCD and DLB participants, as it was not our purpose to compare the CFC to traditional tests that were not designed for these groups. Therefore, SCD and DLB participants who only underwent the MMSE and cognitive battery of the CFC whilst their study partner completed the functional component of the CFC.

### 2.5. Statistical analyses

Statistical analyses were performed using R version 3.5.3 (R Core Team, 2016). Statistical significance was set at *p*<.05. Baseline differences between groups were investigated using Chi-square or Fisher’s exact tests when appropriate, one-way analyses of variance followed by Hochberg’s post-hoc tests, and independent t-tests for measures only available for the MCI and AD groups.

Sensitivity to change over time of the CFC was investigated using linear mixed models (LMM) with random effects for subject (intercept and slope) and center (intercept). All subjects with at least one follow-up assessment available were included in these models. We ran separate models with CFC, CC and AIADLQ scores as dependent variable and time as independent variable (measured on a continuous level). Secondly, we repeated these models whilst adjusting for age, sex, education, diagnosis and the interaction between time and diagnosis. If a significant effect of the time*diagnosis term was found, analyses were repeated stratified per diagnosis. In our sample of MCI and mild AD subjects, separate LMM models adjusting for age, sex and education were also performed with the ADAS-Cog, ADCS-ADL, and CDR-SB as dependent variables and time as independent variables. To control for the different scaling properties and to allow for proper head-to-head comparisons, the ADAS-Cog and CDR-SB total scores were reversed so that higher scores reflected better performance, and subsequently z-transformed using total group baseline means and SD’s of the combined sample of MCI and AD subjects. ADCS-ADL scores were standardized using the same approach.

To compare the sensitivity of the CFC and traditional tests at different time points, *r*^*2*^ effect-sizes of change were calculated from baseline to each follow-up point (3, 6 and 12 months). Effect-sizes were evaluated based on pre-defined cut-offs, with .01 defined as small, .09 as medium and .25 as large effects.

To assess the clinical meaningfulness of observed change on the CFC and traditional tests, separate linear regression analyses were performed for each test in our combined sample of MCI and AD participants. The CFI score obtained at 12 months follow-up was used as dependent variable, and annual change scores were inserted as independent variable while adjusting for age, sex and education.

### Sensitivity analyses

To explore whether change on the CFC was specific to AD-related decline as opposed to change over time in general, we repeated the LMM analyses for the CFC in a subset of amyloid positive participants. Second, differences among study cohorts were explored by repeating the LMM analyses for the CFC and traditional test scores separately for each study center.

## 3. Results

### 3.1. Study sample characteristics

A total of 173 subjects were included (age=71.3±8.5, 42% female, n=14 SCD; n=75 MCI; n=72 AD; n=12 DLB) in the Catch-Cog study, of which n=131 (76%) subjects completed the 12 month assessment (Figure 1). Subjects that withdrew during the study (n=42; age=72.7±8.4, 39% female, n=3 SCD; n=21 MCI; n=15 AD; n=3 DLB) did not differ regarding age, sex, education and clinical severity at baseline from those who completed the study.

**Figure 1.**
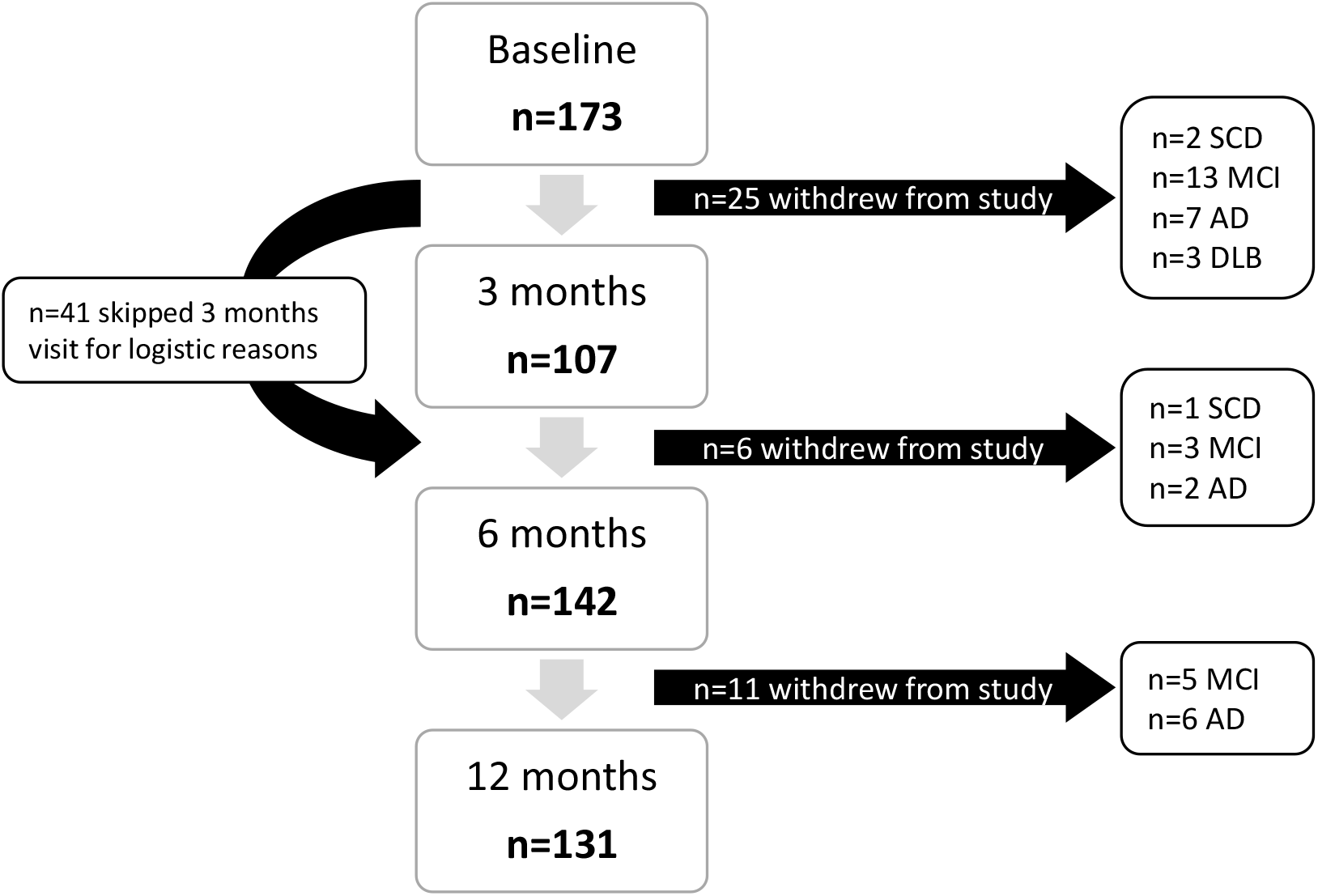
Flow diagram providing an overview of the sample-size at each time-point.

A number of n=148 participants (86%) had at least one follow-up available and were included in the current study. Table 1 presents the baseline characteristics for this sample as well as separately for each clinical group, revealing that groups did not differ regarding age, sex and education. Mean baseline CFC scores differed among groups (SCD=.91±.61; MCI=.28 ±.50; AD=−.34±.65; DLB=−.51±.75, *F*=25.84, *p*<.001), with post-hoc comparisons showing a significant higher score for SCD compared to MCI, as well as for MCI compared to AD and DLB dementia (Table 1). A similar pattern was observed on the traditional tests, except that no significant differences were found in the ADAS-Cog scores between the MCI and AD group (Table 1).

**Table 1.**
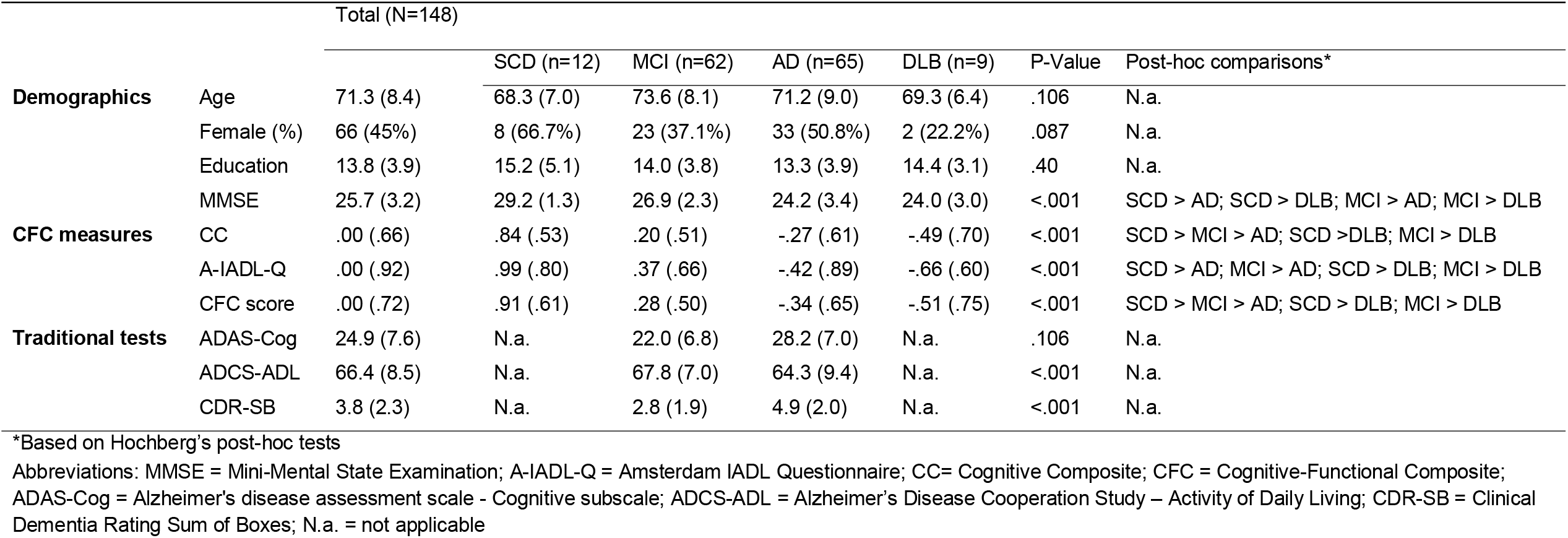
Demographic and clinical characteristics at baseline, for the total included sample (N=148) as well as separately for each clinical group.

### 3.2. Sensitivity to change over time of the CFC in the total sample

Overall, CFC scores declined over time (*β*=−.15, 95%CI [−.10 – −20], *p<*.001), and we found that this association was independent of age, sex, education and diagnosis at baseline (corrected *β*=−.16, 95%CI [−.10 – −22], *p<*.001). However, a significant time*diagnosis interaction was found (*β=−.18, p*<.001), and therefore analyses were repeated stratified per clinical group. The results are presented in Figure 2, showing that decline on the CFC was only significant in the mild AD group (*β*=−.25, 95% CI [−.33 – −18], *p<*.001). A steeper decline was observed in DLB, however, this effect did not reach statistical significance (*β*=−.47, *p*=.096).

**Figure 2.**
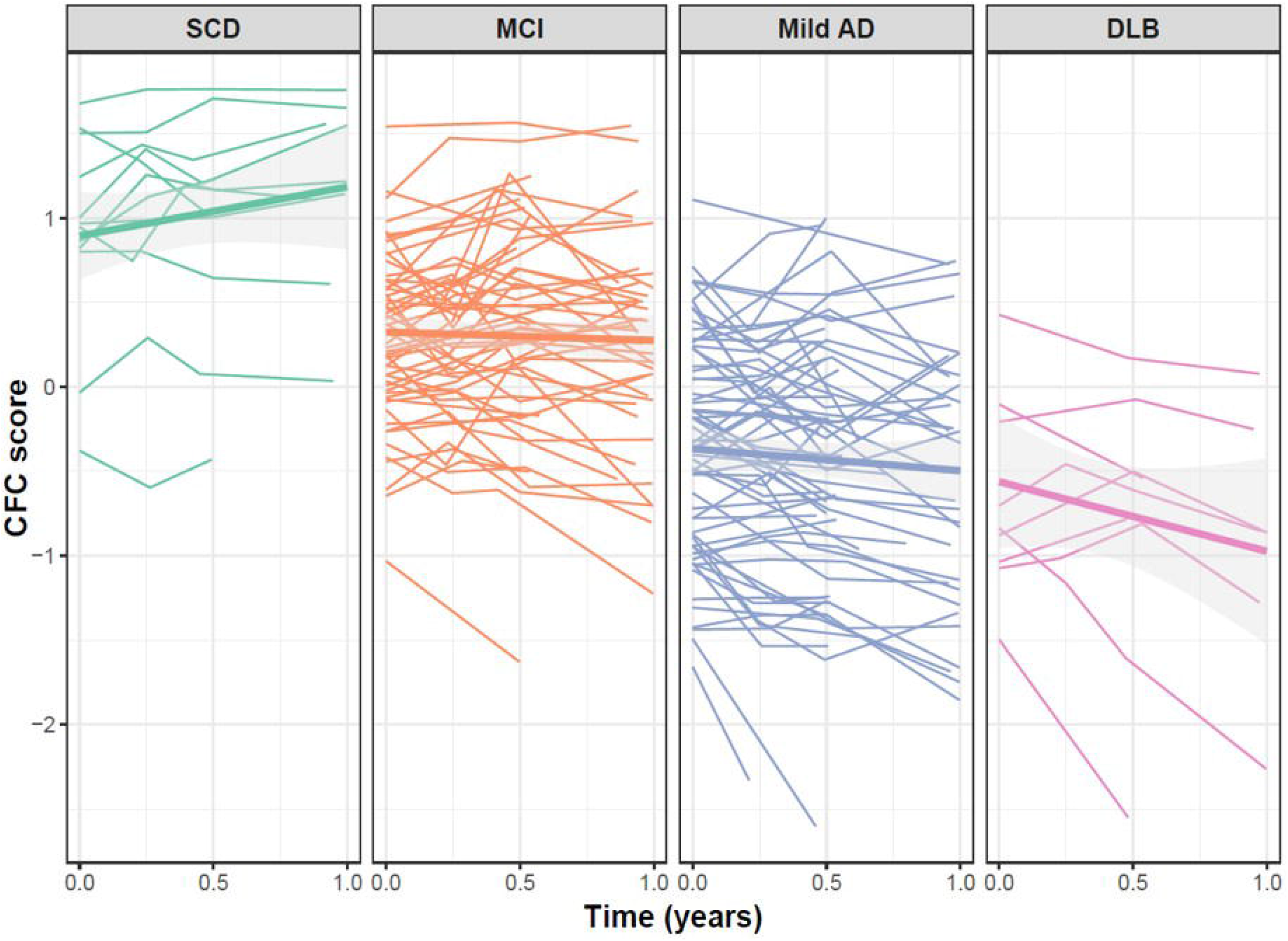
Annual change on the CFC, separately for each clinical group.

### 3.3. Comparison between the CFC and traditional measures in MCI and mild AD

Figure 3 displays change on the CFC components and the traditional tests adjusted for age, sex and education, separately for the MCI and AD groups. Supplementary Table 1 and 2 show the corresponding regression coefficients. We found a significant decline in A-IADL-Q score in MCI (*β*=−.15, *p*=.03), whereas none of the traditional measures declined in MCI (Supplementary Table 2).

**Figure 3.**
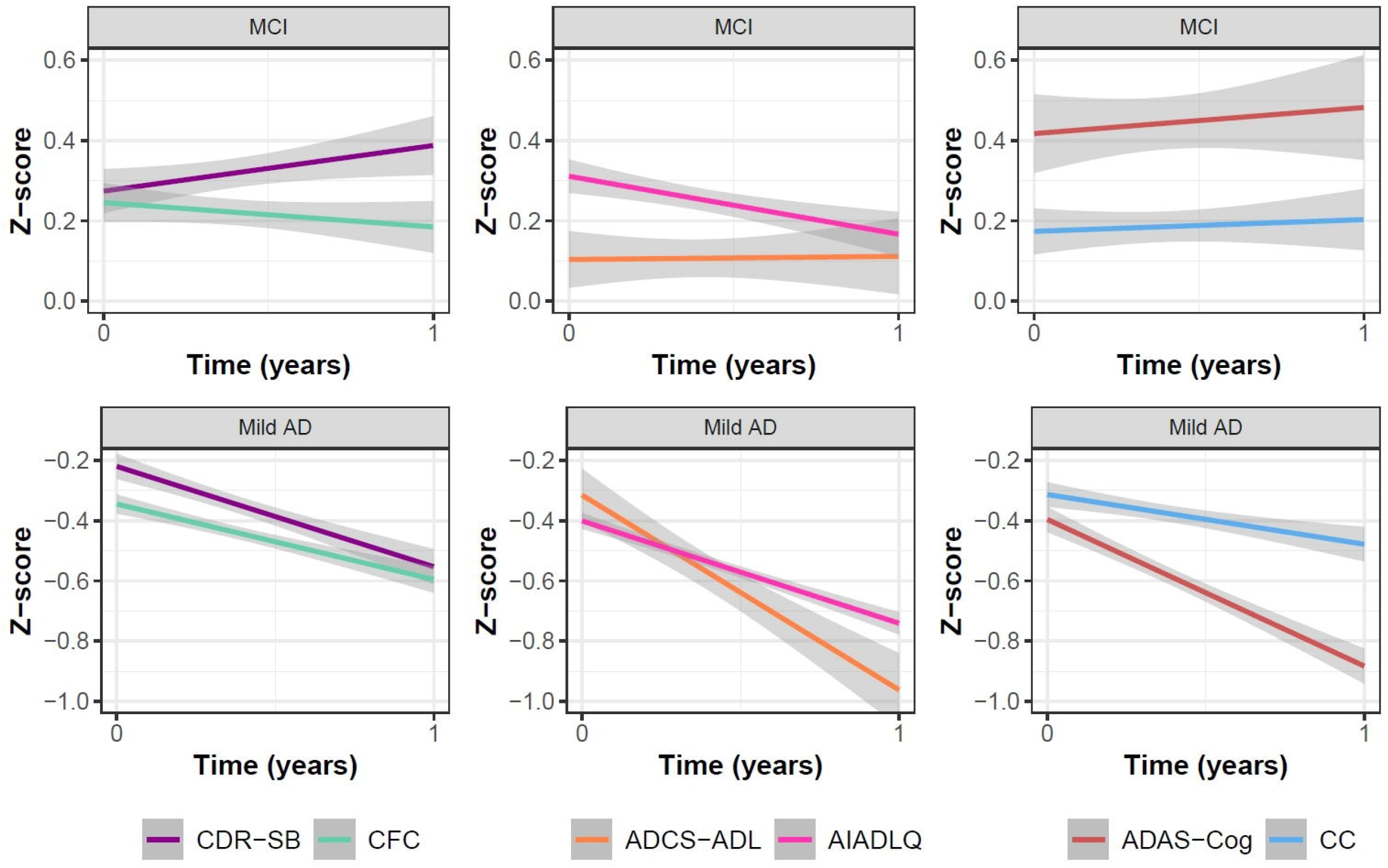
Annual decline (corrected for age, sex and education) on the CFC measures versus traditional tests, separately in MCI and AD.

All CFC scores and traditional tests scores declined over one year in AD (Figure 3, Supplementary Table 1 and 2). Effect-sizes of change at all follow-up time-points on the CFC and traditional tests are presented in Figure 4. The CFC showed a small-to-medium effect after 3 months (*r*^*2*^=.08), a medium effect after 6 months (*r*^*2*^=.12), and a large effect (*r*^2^=.42) after 12 months, thereby exhibiting greater change than the CDR-SB all follow-up time points (*r*^2^ ranging from .02–.12).

**Figure 4.**
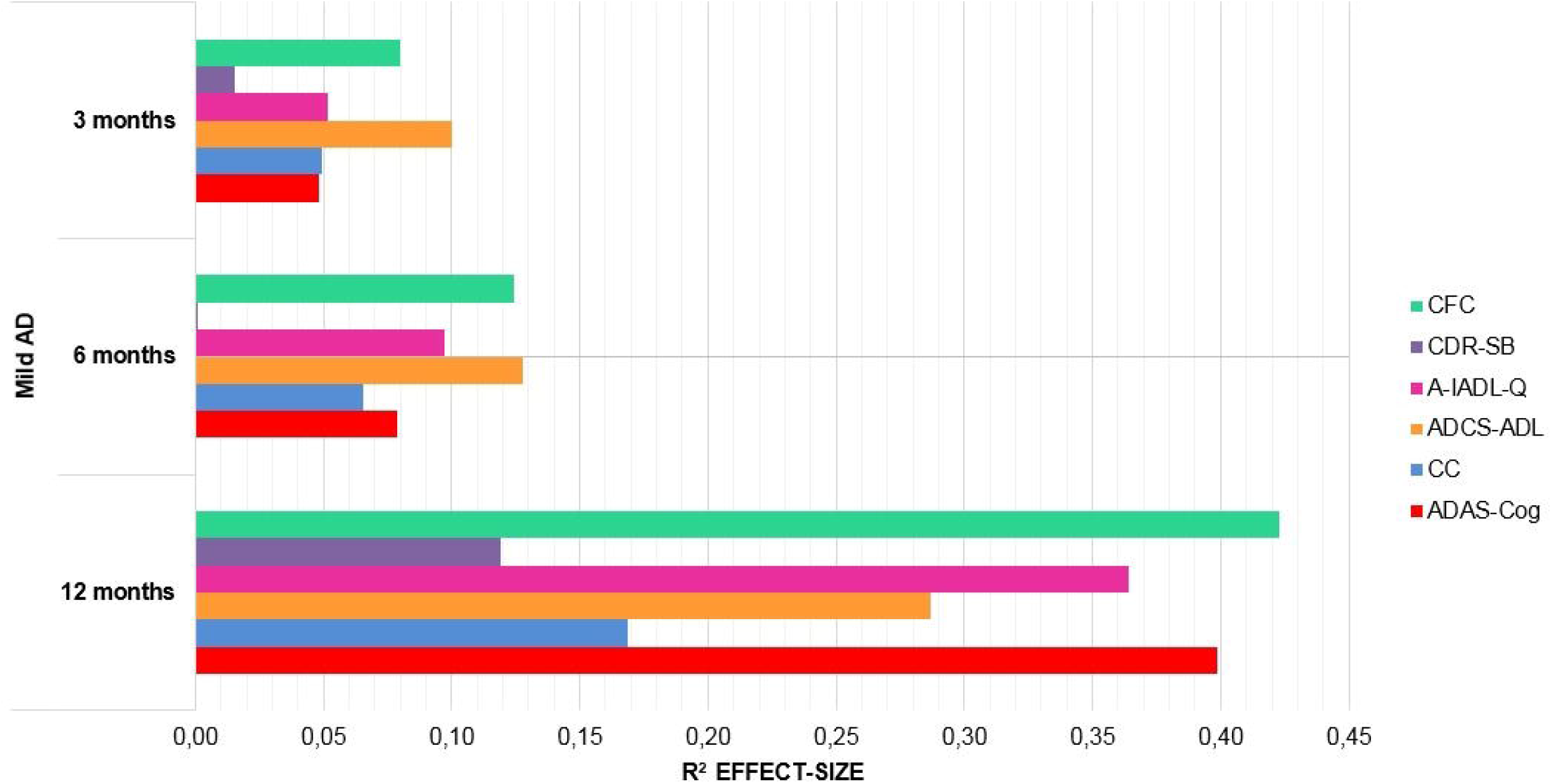
Effect-sizes of the CFC measures and traditional tests at all follow-up time points.

### 3.4. Association with an anchor measure of clinical progression in MCI and mild AD

Figure 5 shows the associations between the CFI score obtained at 12 month follow-up and annual decline on the CFC, CDR-SB, ADCS-ADL and ADAS-Cog in our combined sample of MCI and mild AD subjects (n=127). Linear regression analyses showed that decline on the CFC was significantly associated with decline in cognitive functioning as reported on the CFI (*β*=.38, 95% CI [.20 – .56], *p*<.001). Among the traditional tests, only decline on the ADCS-ADL was significantly related to the CFI score at 12 month follow-up (*β*=.26, 95% CI [.07− .45], *p*=.009).

**Figure 5:**
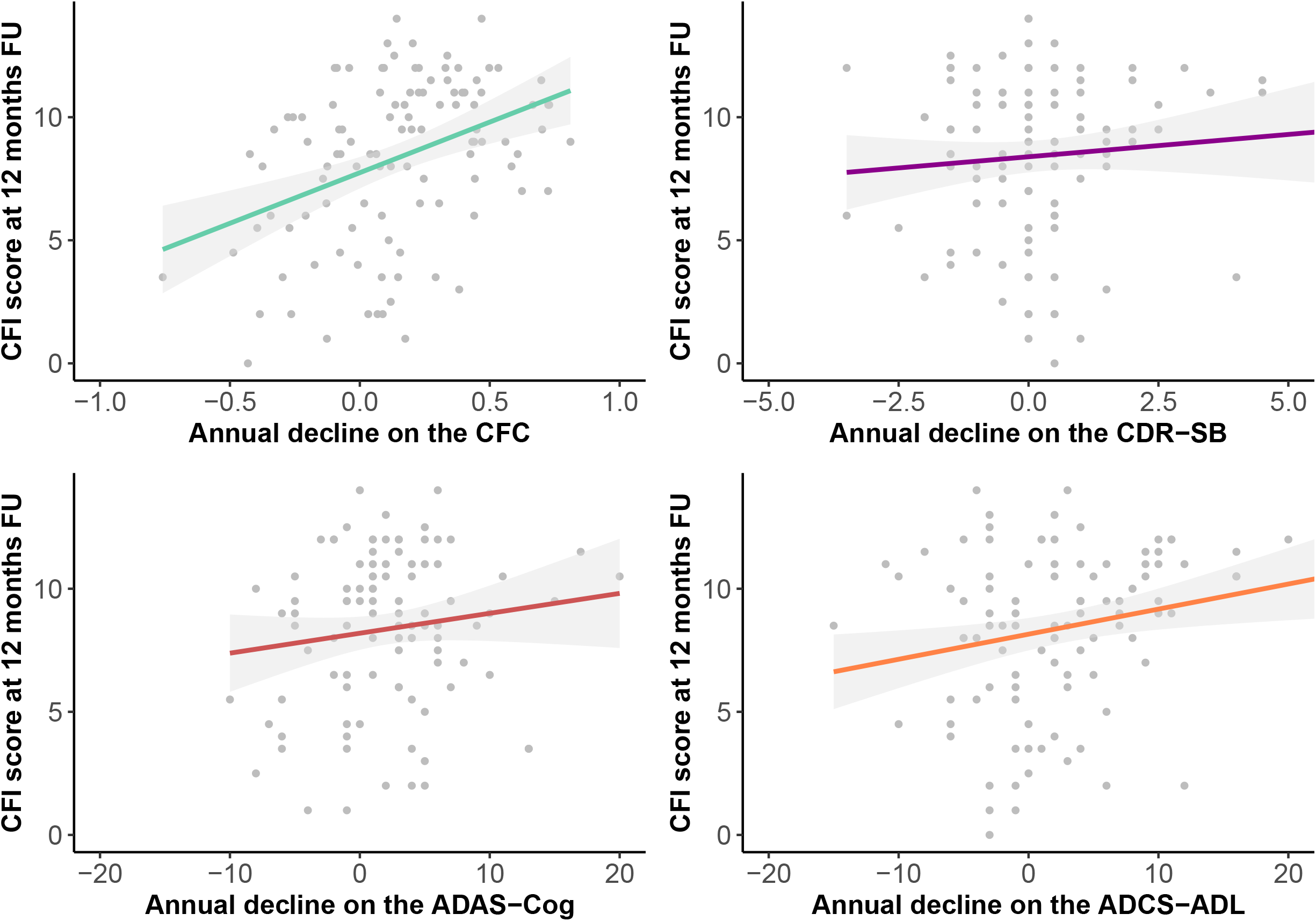
Associations between CFI score at 12 months follow-up and annual decline in CFC, CDR-SB, ADAS-Cog and ADCS-ADL scores. N.B. All x-axis scales represent annual change scores with positive scores reflecting decline compared to baseline.

### 3.5. Sensitivity analyses

Sensitivity analyses in amyloid positive participants (n=37; n=1 SCD, n=7 MCI and n=29 dementia) showed that CFC scores declined over time (*β*= −.21, 95% CI [−.11 – −.33], *p<*.001), with decline observed on both the cognitive and functional component (CC score: *β*= −.16, 95% CI [−.29 – −.04], *p<*.001; A-IADL-Q score: *β*= −.30, 95% CI [−.47 – −.14], *p<*.001).

Supplementary Table 3 shows the regression coefficients obtained from LMM stratified per study center. These results show that CFC scores declined over time in the AC, EMC and EDI cohorts, whereas a slight improvement on the CFC (*β*=.11, *p*=.049) was observed in the UMCG cohort that seemed driven by the cognitive component (*β*=.19, *p<*.001). This was in agreement with results on the traditional tests, as an improvement on the ADAS-Cog was also observed in UMCG cohort (*β*=.39, *p*=.003).

## 4. Discussion

In this longitudinal construct validation study, we demonstrated that the CFC is sensitive to clinical progression. In MCI, the functional component of the CFC detected decline over one year, whereas the CDR-SB, ADAS-Cog and ADCS-ADL failed to do so. In mild AD dementia, the CFC captured decline over one year, and effect-sizes of change suggested greater sensitivity than the traditional measures within one year (i.e. after 3 and 6 months). Furthermore, annual change on the CFC was associated with informant-reports of cognitive decline.

Worldwide, researchers have addressed the need for outcome measures that are capable of detecting clinically meaningful change in early AD [14, 41]. Regulatory agencies have further elevated this importance, as their guidelines state that evidence of clinically meaningful change is required for approval of novel therapeutic interventions [15, 16]. Previous studies on the CFC already demonstrated its good test-retest reliability, feasibility of use, construct validity, and suitability for the target population (i.e. MCI and mild dementia due to AD) [27, 28, 42]. Additionally, separate studies on the cognitive and functional component previously showed their sensitivity to change over time [23, 43]. By performing an independent longitudinal validation of the CFC, the current study provides further evidence the CFC meets the requirements for a clinically meaningful outcome measure in early clinical stages of AD.

The current study results bear crucial implications for AD clinical trials, since the CDR-SB is currently still widely-applied as primary clinical endpoint of efficacy. Previous studies have already indicated limitations of the CDR-SB as outcome measure of change, relating to its poor inter-rater reliability [21] and ceiling effects in scoring in MCI and mild dementia [28]. Our current study findings suggest that the CFC could offer advantages over the use of the CDR-SB as measure to evaluate clinical progression, as it provides a concise and objective measure of clinically meaningful cognitive decline.

When considering other recently developed composites for disease progression in AD, it is important to consider that CDR performance is also a key component of the recently employed Alzheimer’s Disease Composite Score (ADCOMS) [44]. However, this measure has been statistically-derived, and its clinical meaningfulness has not been demonstrated yet. The same holds true for proposed cognitive composites [45,46], which have been developed to detect change in preclinical AD. Since those composites do not include a functional component, they are probably less useful to track clinical progression in MCI and mild dementia stages, in which evolving functional impairment plays a key role [42].

Comparisons between the CFC subcomponents and traditional cognitive and functional tests revealed that the A-IADL-Q already captured decline in MCI, whereas the ADCS-ADL did not. This implies that the A-IADL-Q is more focused on those activities that are prone to decline in earlier clinical stages [42]. Interestingly, comparable sensitivity to change was observed for the CC and ADAS-Cog. This could be explained by the fact that those measures partly overlap, and that changes on the ADAS-Cog score seemed driven by the three memory subtests that are also included in the CC. This is line with previous studies showing that the ADAS-Cog subtests that focus on praxis, language and confrontation were found to be insensitive to change in MCI and mild dementia [7]. As such, the CC can be considered a more concise measure, as it has a shorter administration time and focuses on the cognitive domains that are vulnerable to early cognitive decline [47].

Our finding that the functional component of the CFC detected clinical progression whereas the cognitive component did not, may seem counterintuitive as the assumed clinical trajectory of AD entails that cognitive impairment induces and thereby precedes functional impairment [48]. However, our findings do not argue against this conceptual understanding of cognitive impairment preceding functional change, but rather imply that existing paper-and-pencil cognitive tests may not provide the right tools to capture subtle cognitive decline. This is in line with previous studies that pointed towards the limited sensitivity of existing cognitive tests in early clinical stages of AD [49, 50]. A functional measure on the other hand, may be capable of capturing meaningful decline as reflected by increasing difficulties in complex activities of daily living.

There are some limitations that should be considered. First, our results might have been biased by heterogeneity in our sample due to differences in recruitment strategies employed across the centers. The majority of the UMCG cohort included community-based SCD and MCI participants, who are presumed to be at less risk for developing dementia as compared to those recruited in a memory clinic setting [51]. It is therefore likely that those community-based participants showed less progression in cognition and function over time, as also reflected by our sensitivity analyses after stratification by study center. As such, our main analyses on the CFC’s sensitivity to clinical progression may have been underestimated, especially in the MCI group. The fact that those community-based individuals did also not decline on the traditional tests indicates that this limitation was not specific for the CFC. Second, the sample-size of the SCD and DLB groups were relatively small and thus the power to detect statistically significant was limited. Hence, no strong interferences can yet be drawn for the utility of the CFC in these groups. Third, it could be argued that a follow-up period of one year is relatively short to observe evident cognitive decline in individuals with MCI or mild AD. However, we think it is a relevant timeframe as it corresponds to regular follow-up periods in clinical practice and clinical trial designs, and therefore it is crucial to know whether the CFC can capture clinical changes within this timeframe. Finally, it should be noted that the same rater completed all assessments within one visit and was therefore not blind for the cognitive assessments when performing the study partner interview, and that our CDR-SB assessment was slightly different than in other studies. However, we do not think that this has affected our CDR-SB scores, nor our comparisons between the CDR-SB and CFC.

Strengths of this study include our study design that enabled us to perform an independent validation of the CFC. Furthermore, the direct comparisons between the CFC and traditional measures is an important and unique aspect of this study. Additionally, our comparison with an anchor measure of everyday functioning strengthened the clinical meaningfulness of our findings. This additional investigation has been founded on the concern that statistically significant effects observed on cognitive tests do not self-evidently demonstrate clinically meaningful effects [9]. It should be acknowledged that our approach of establishing clinical meaningfulness has limitations, and that more sophisticated, qualitative methods involving patients and experts focus groups exist [52]. However, we think the current study provides a first step to assess the CFC’s clinical meaningfulness. Finally, sensitivity analyses in the amyloid-positive group enabled us to explore whether the CFC would be sensitive to AD-specific decline. However, these findings should be interpreted with caution due to the relatively small sample-size of participants with biomarker data available, and thus need to be replicated in a larger sample. Altogether, these aspects will likely enhance future implementation of the CFC in both AD research and clinical practice.

Future directions include the optimization of the CFC’s sensitivity to change in MCI, for example by exploring different weights for the CFC components to create a more sensitive score. Additionally, applying IRT scoring to the cognitive component might yield more precise measurement and thereby aid the detection of change over time [53]. IRT has already been applied in the scoring of the A-IADL-Q, which showed the highest sensitivity to change. Furthermore, IRT would enable us to use anchor-based bookmarking methods to determine the minimal important change, which could further establish the clinical meaningfulness of the CFC. Additionally, it would be interesting to investigate the sensitivity of the CFC beyond one year of follow-up, with a particular focus on the SCD and MCI groups. This would allow us to investigate whether the sensitivity of the CFC could be improved, and whether the CFC could predict conversion to dementia. To further facilitate the implementation of the CFC in clinical practice and research, it would be relevant to develop norms based on CFC performance in cognitively normal individuals. Lastly, it would be interesting to further investigate the utility of CFC to measures progression in DLB, given the increasing number of DLB clinical trials [54] and the limited understanding on the clinical course of DLB so far [55].

In conclusion, the current study provides evidence that the CFC yields an efficient and clinically meaningful measure of disease progression, and thereby has the potential to serve as efficacy endpoint for use in AD clinical trials [15, 16]. By providing a concise measure of clinically meaningful cognitive decline, the CFC could contribute to the monitoring of disease progression, which is of relevance for both research and clinical practice.

## Data Availability

Data is available upon reasonable request.

## Acknowledgements

The authors would like to thank Philippe Lee Meeuw Kjoe, Mandy Ter Haar, Larissa Masselink, Judith Meurs, Mieke Geertsma, Nina Schimmel, Ilya de Groot, Judy van Hemmen, Kate Forsyth, Sarah Gregory, Neil Fullerton, Clare Dolan and Matthew Hunter for their help with the data collection. Additionally, we would like to acknowledge Stichting Buytentwist for their support. Research of the Alzheimer Center Amsterdam is part of the neurodegeneration research program of Amsterdam Neuroscience. The Alzheimer Center Amsterdam is supported by Alzheimer Nederland and Stichting VUmc Fonds. The present study is supported by a grant from Memorabel (grant no. 733050205), which is the research program of the Dutch Deltaplan for Dementia.

## List of Abbreviations

AC: Alzheimer Center Amsterdam
AD: Alzheimer’s Disease
ADAS-Cog: Alzheimer’s Disease Assessment Scale - Cognitive subscale
ADCOMS: Alzheimer’s Disease Composite Score
ADCS-ADL: Alzheimer’s Disease Cooperative Study - Activities of Daily Living Inventory
AES: Apathy Evaluation Scale
A-IADL-Q: Amsterdam IADL Questionnaire
Catch-Cog: Capturing Changes in Cognition
CC: Cognitive Composite
CDR-SB: Clinical Dementia Rating scale – Sum of Boxes
CFC: Cognitive Functional Composite
CFI: Cognitive Function Instrument
CFT: Category Fluency Test
COWAT: Controlled Oral Word Association Test
DLB: Dementia with Lewy Bodies
DSST: Digit Symbol Substitution Test
EDI: Centre for Dementia Prevention, Edinburgh
EM: Episodic memory
EMA: European Medicine Agency
EMC: Erasmus Medical Center
EF: Executive functioning
FDA: Food and Drug Administration
IADL: Instrumental Activities of Daily Living
IRT: Item Response Theory
LMM: Linear Mixed Models
MCI: Mild Cognitive Impairment
MMSE: Mini-Mental State Examination
SCD: Subjective Cognitive Decline
UMCG: University Medical Center Groningen
WM: Working Memory.

